# PDK4-Mediated VDAC2 Lactylation Links Glycolysis to Autophagy Failure in Septic Cardiomyopathy

**DOI:** 10.1101/2025.09.22.25336402

**Authors:** Nuo Xu, Lu Gan, Yuhan Feng, Shirong Chen, Junli Luo, Jinzi Chen, Jiayi Wu, Yili Wang, Liping Huang, Shuangjiao Chao, Zehong Huang, Jiawei Ni, Kexuan Liu, Cai Li

**Affiliations:** Department of Anesthesiology, Nanfang Hospital, Southern Medical University, Guangzhou, 1838 Guangzhou Avenue North, Guangzhou, 510515, China; Guangdong Provincial Key Laboratory of Precision Anesthesia and Perioperative Organ Protection, Guangzhou, 510515, China

**Keywords:** Sepsis-induced cardiomyopathy, PDK4, Lactylation, VDAC2, Autophagy

## Abstract

**Background:** Elevated lactate is a critical prognostic biomarker in sepsis, strongly associated with increased mortality. Sepsis-induced cardiomyopathy (SICM) is a major contributor to poor outcomes in septic patients, yet the mechanisms linking lactate elevation to SICM pathogenesis remain incompletely understood.

**Methods:** We combined experimental murine models of sepsis with patient cohort analyses to investigate the metabolic and molecular mechanisms underlying SICM. Glycolytic flux, lactate production, and protein lactylation were assessed using metabolomics, proteomics, and site-directed mutagenesis. The clinical relevance of pyruvate dehydrogenase kinase 4 (PDK4) was evaluated by correlating serum levels with lactate and cardiac injury biomarkers in septic patients.

**Results:** In experimental SICM, PDK4 upregulation enhanced glycolysis and promoted lactate accumulation. Elevated lactate induced lactylation of voltage-dependent anion channel 2 (VDAC2), specifically at lysine 75 (K75). Mechanistically, VDAC2 K75 lactylation disrupted its interaction with neighbor of BRCA1 gene 1 (NBR1), suppressing cardiomyocyte autophagy and exacerbating myocardial injury. Clinically, serum PDK4 levels positively correlated with lactate and cardiac injury markers, showing strong predictive value for SICM (AUC = 0.8516).

**Conclusions:** Our findings identify a pathogenic axis whereby PDK4-driven metabolic reprogramming promotes VDAC2 lactylation, impairs cardiomyocyte autophagy, and accelerates cardiac dysfunction in SICM. Targeting the PDK4–lactate–VDAC2 pathway may represent a novel therapeutic strategy for improving outcomes in septic cardiomyopathy.

**Clinical Perspective:** *What Is New?:*

- Elevated lactate is strongly associated with mortality in sepsis, but its mechanistic contribution to SICM was unknown.
- This study identifies a pathogenic axis in which PDK4 promotes glycolysis-dependent lactate accumulation, driving VDAC2 lactylation at lysine 75.
- VDAC2 lactylation disrupts interaction with NBR1, impairs autophagy, and exacerbates cardiac injury.
- In septic patients, serum PDK4 levels correlate with lactate and cardiac injury markers, showing predictive value for SICM.

*What Are the Clinical Implications?:*

- Serum PDK4 may serve as a potential biomarker for the identification of patients with sepsis-induced cardiomyopathy.
- Targeting the PDK4–lactate–VDAC2 pathway represents a promising therapeutic strategy to restore autophagy and attenuate SICM.
- These findings link metabolic reprogramming to SICM pathogenesis and provide a translational framework for intervention.

## Introduction

Sepsis is a life-threatening condition characterized by a dysregulated host response to infection, leading to excessive cellular stress, immune dysfunction, and ultimately, multiple organ failure ^1, 2^. Among its complications, sepsis-induced cardiomyopathy (SICM) stands as one of the most common and fatal, representing a major contributor to sepsis-related mortality ^3, 4^. Elevated blood lactate levels are strongly associated with a poor prognosis in septic patients, while effective lactate clearance significantly improves survival ^5–8^. Although our previous findings demonstrated a positive correlation between lactate levels and myocardial injury markers in septic patients, the underlying mechanisms remain poorly understood.

Lactate is recognized as a key metabolite produced via glycolytic reprogramming during sepsis. This metabolic shift is regulated by critical proteins, including hypoxia-inducible factor 1-α (HIF-1α) ^9^, pyruvate kinase M2 (PKM2) ^10^, and hexokinase 2 (HK2) ^11^. Among these, pyruvate dehydrogenase kinase 4 (PDK4) plays a pivotal role in regulating cellular energy balance by modulating the glycolysis–oxidative phosphorylation axis ^12^. PDK4 phosphorylates and inhibits the pyruvate dehydrogenase complex (PDC), thereby diverting pyruvate away from mitochondrial oxidation towards lactate-producing glycolysis and promoting lactate accumulation ^13^. PDK4 is highly expressed in cardiac and hepatic tissues, and existing evidence implicates it in SICM pathogenesis through mechanisms involving mitochondrial dysfunction ^14, 15^. However, the precise molecular pathways require further elucidation.

Recent studies have revealed a novel role for lactate beyond its metabolic function. Traditionally viewed as a hypoxia byproduct, lactate now emerges as a substrate for protein lactylation—a recently discovered post-translational modification ^16^. Protein lactylation has been shown to modulate various cellular functions, including macrophage polarization ^11^, inflammatory responses ^17, 18^, and cell death ^18^, and has been implicated in sepsis-related injuries in the lungs ^19, 20^ and kidneys ^21–23^. Preliminary data also suggest that protein lactylation may impair cardiac function in sepsis; however, the involvement of other lactylated proteins and their downstream effects remains largely unexplored.

Mitochondrial quality control, particularly through autophagy, is critical for cardiomyocyte survival, and its dysfunction is a key pathological mechanism in sepsis-induced myocardial injury ^24–26^. Selective autophagy receptors such as the neighbor of BRCA1 gene 1 (NBR1) recognize ubiquitinated cargo and facilitate their degradation by linking them to LC3-positive autophagosomes ^27, 28^. The voltage-dependent anion channel 2 (VDAC2), a β-barrel transmembrane protein located on the mitochondrial outer membrane, plays a key role in metabolite transport and mitochondrial function ^29–31^. Previous studies have shown that VDAC2 regulates cardiac function via calcium modulation ^30^ and that it also participates in autophagy during ovarian development ^32^. In light of the crucial positioning of VDAC2 on the outer mitochondrial membrane and its function in autophagy, we hypothesized that it could be an important sepsis-induced lactylation target, therefore linking metabolic stress to mitochondrial quality control failure. However, the regulatory crosstalk between VDAC2 and NBR1, especially during sepsis, remains unknown.

In this study, we explored the role and underlying mechanisms of lactate and its upstream regulator PDK4 in the development of SICM. Our findings reveal that PDK4 promotes lactate accumulation by enhancing glycolytic activity. The accumulated lactate facilitates protein lactylation, which in turn aggravates myocardial injury and increases mortality in septic mice. Importantly, we identified that VDAC2 lactylation inhibits autophagic flux by attenuating its interaction with NBR1, a key mechanism driving cardiac dysfunction in sepsis. Collectively, our results delineate a novel pathogenic cascade in SICM, consisting of PDK4-mediated lactate production, subsequent VDAC2 lactylation, autophagy inhibition, and ultimately, myocardial injury. These findings suggest that inhibiting PDK4 or lactate production, or VDAC2 lactylation may serve as potential therapeutic strategies for preventing and treating SICM.

## Methods

Detailed Methods are available in the Supplemental Material and Methods. All animal experimental procedures were carried out in accordance with the National Institutes of Health guidelines and were approved by the local Animal Care and Use Committee of the Nanfang Hospital of Southern Medical University (approval number: IACUC-LAC-20220701-002). The acquisition and use of serum from sepsis patients roved by the Ethic Committee of Nanfang Hospital (Ethics approval number: NFEC-2025-382).

### Quantification and Statistical Analysis

All statistical analyses were performed using GraphPad Prism 9.0 software (GraphPad Software, Inc., LaJolla, CA, USA). Quantitative data are presented as the means ±standard deviation. For all animal and cell experiments with a small sample size (*n*≤8), the Shapiro–Wilk test was used to test for normality with a threshold of 0.05. For data with normal distribution, the Student’s *t*-test was used to analyze the significance of differences between two means; to compare several groups, one-way ANOVA with Tukey’s or Dunnett’s multiple-comparison test was used when variances were equal, while One-way ANOVA with Welch’s correction was used when variances were unequal; correlation analysis was performed using Pearson correlation.

For non-normally distributed data, the Mann–Whitney or Kruskal–Wallis test was used to test the significance of differences, and correlation analysis was per formed using Spearman correlation. Survival analysis in mice was performed using the LOG-RANK test. A *p* value < 0.05 were considered statistically significant.

## Results

### PDK4 is significantly elevated during SICM and drives cardiomyocyte glycolysis to promote lactate accumulation

In both *in vivo* and *in vitro* models of SICM, serum lactate levels were significantly elevated (Figure 1A, Figure S1A). To explore the upstream mechanism, we measured key metabolic enzymes including Pyruvate Dehydrogenase Kinase 1-4 (PDK1-4), Hexokinase 2 (HK2), Pyruvate Kinase M2 Isozyme (PKM2), 6-Phosphofructo-2-Kinase/Fructose-2,6-Biphosphatase 3 (PFKFB3) and Lactate Dehydrogenase A (LDHA). Among these, only the levels of PDK4 and PFKFB3 were significantly increased (Figure 1B). Proteomic sequencing confirmed the PDK4 elevation (Figure 1C), a finding further validated in LPS-treated H9C2 cells and mouse cardiac tissue (Figure 1D-1E, Figure S1B-C). These findings indicate a potentially critical role of PDK4 in SICM.

**Figure 1.**
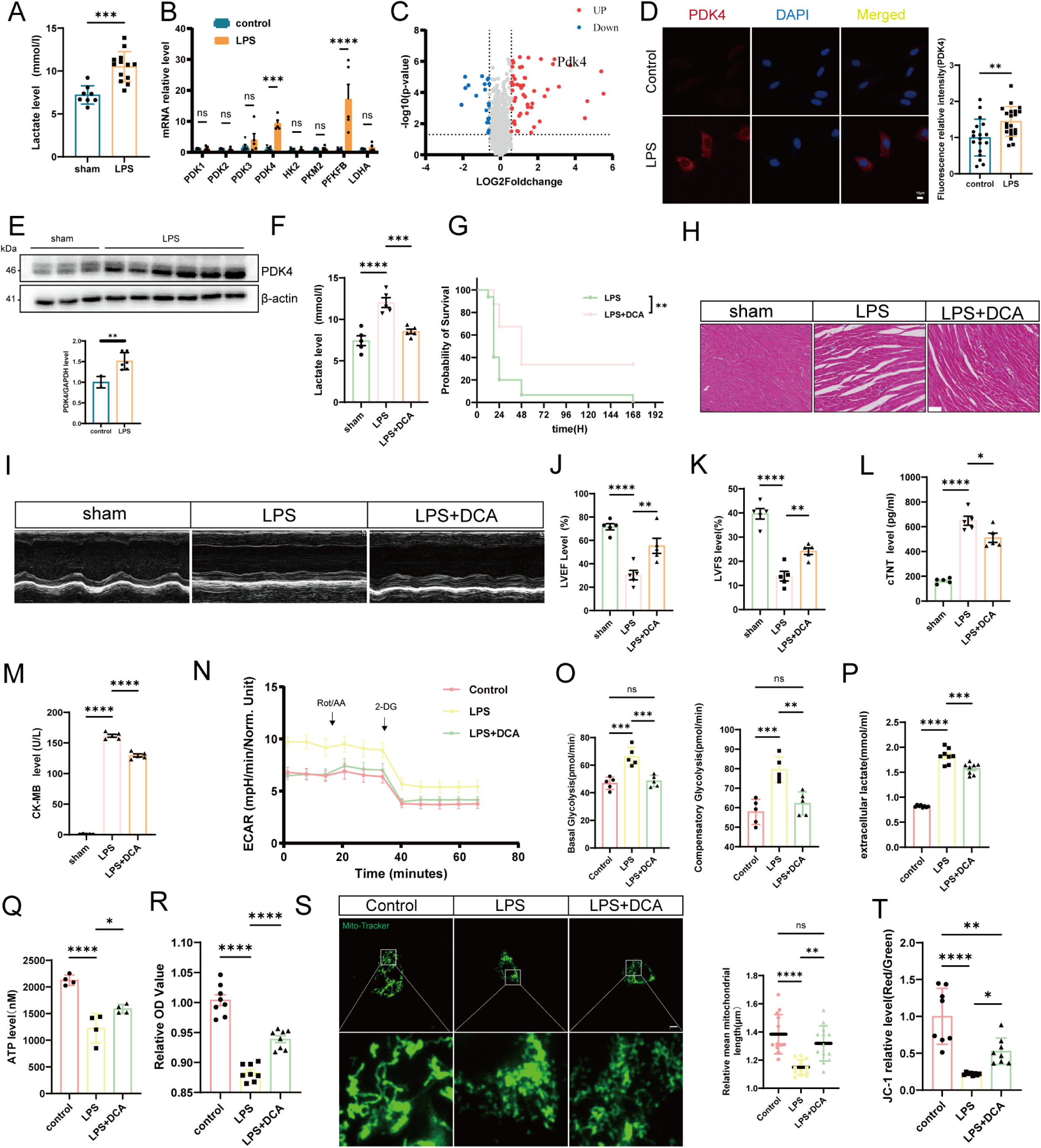
PDK4 is significantly elevated during SICM and drives cardiomyocyte glycolysis to promote lactate accumulation. (A) Serum lactate concentrations in Control and LPS-treated mice (Control n=8, LPS n=13). (B) mRNA expression levels of PDK1, PDK2, PDK3, PDK4, HK2, PKM2, PFKB3, and LDHA in myocardial tissues of Control and LPS-treated mice (Control n=4; LPS n=5). (C) Volcano plot differential protein expression in murine myocardial tissue, with only PDK4 highlighted (n=3). (D) Representative immunofluorescence images of PDK4 staining in H9C2 cells (scale bars: 10 μm). (E) Western blot analysis of PDK4 protein in mouse myocardial tissues (sham n=3, LPS n=6). (F) Serum lactate levels in sham, LPS, and LPS+DCA-treated mice (n=5). (G) Survival analysis of LPS, and LPS+DCA-treated mice (n=16). (H) Representative HE-stained images of heart tissues (scale bars: 100 μm). (I)Representative M-mode echocardiograms of mice treated with sham,LPS or LPS+DCA. (J, K) LVEF and LVFS in Control, LPS, and LPS+DCA-treated mice (n=5). (L, M) Myocardial injury marker (CK-MB, cTnT) levels in Control, LPS, and LPS+DCA-treated groups (n=5). (N-O) Seahorse metabolic analysis (extracellular acidification rate, ECAR) of H9C2 cardiomyocytes treated with control, LPS, or LPS combined with DCA (n=5). (P) Lactate content in the supernatant of H9C2 cells across different treatment groups (n=8). (Q) ATP content in H9C2 cells across different treatment groups (n=4). (R) Cell viability of different treatment groups (n=8). (S) Mitochondrial morphology in H9C2 cells visualized by MitoTracker-Green staining and analyzed using the MiNA plugin in ImageJ software (scale bar: 10 μm). (T) Mitochondrial membrane potential in H9C2 cells treated with control, LPS, or LPS combined with DCA. Data are mean ± SD. The statistical significance of the differences between groups was determined by unpaired two-tailed Student’s t test (A,E) or Two-way ANOVA with Bonferroni’s multiple comparisons test(B) or Mann-Whitney test (D) or Brown-Forsythe and Welch ANOVA tests (M) or Log-rank (Mantel-Cox) test (G) or one-way ANOVA with Dunnett’s post hoc analysis (F, J, K, L,O,P,Q,R,S and T) (n.s., not significant; *p < 0.05, **p < 0.01 versus sham or control or sham or LPS).

To validate PDK4’s function, we first determined its relationship with lactate accumulation. Pharmacological inhibition (DCA) or gene knockout (AAV) of *PDK4* significantly reduced serum lactate (Figure 1F, Figure S1D) and increased mouse survival (Figure 1G).

Consistent with reduced lactate, both treatment protocols significantly mitigated SICM. Pathologically, this was characterized by reduced interstitial expansion and cardiomyocyte damage (Figure 1H and Figure S1E). Functionally, cardiac parameters—fractional shortening (FS) and left ventricular ejection fraction (LVEF)—were significantly enhanced in these mice (Figure 1I-1K, Figure S1F-S1H). Serum indicators of cardiac damage (cardiac troponin T (cTnT), creatine kinase-MB (CK-MB) were also reduced compared to the septic group (Fig. 1L-1M, S1I-S1J), as seen in *PDK4*-knockdown H9C2 cells (siRNA) (Figure S1K-S1L). Western blot confirmed PDK4 to be stably knocked down (Figure S1M).

At the cellular level, these results were due to the role of PDK4 in glycolysis. LPS treatment increased the level of extracellular acidification rate (ECAR), an action that was reversed by PDK4 inhibition or knockdown (Figure 1N-1O, Figure S1N-S1P) in H9C2 cells. Extracellular lactate level went in the same direction (Figure 1P and Figure S1Q), as did cell ATP level (Figure 1Q), with cell viability enhanced (Figure 1R, Figure S1R). Furthermore, mitochondrial length, which decreased following LPS treatment, was partially restored by PDK4 inhibition or knockdown (Figure 1S, Figure S1S). JC-1 staining revealed similar reversal patterns for mitochondrial membrane potential (Figure 1T, Figure S1T). Collectively, these results demonstrate that PDK4-dependent glycolysis, which is followed by lactate accumulation, harms mitochondria and that its inhibition diminishes SICM.

### PDK4 and lactate may serve as biomarkers for SICM in patients with sepsis

To translate our findings from the animal model, we next investigated the clinical relevance of PDK4 and lactate in a cohort of septic patients. PDK4 is highly conserved (>90% amino acid identity between humans and mice), and our prior work linked elevated PDK4 expression to septic myocardial injury. We therefore hypothesized that serum PDK4 could serve as a biomarker. Fifty-five septic patients were enrolled: 13 patients in the Sepsis-induced non-cardiomyopathy (SINCM) group and 42 patients in the SICM group. With the exception of age, the SINCM and SICM groups showed no significant differences in baseline characteristics, including gender distribution and Sepsis-related Organ Failure Assessment (SOFA) scores. Serum lactate, high-sensitivity cardiac troponin T (hs-TnT), and total ICU length of stay were significantly higher in the SICM group (Table S1).

Both PDK4 levels and lactate levels were significantly elevated in the SICM group (Figure 2A-2B) and showed a positive correlation with each other (Figure 2C). PDK4 correlated positively with hs-TnT and B-type natriuretic peptide (BNP) (Figure 2D–2E). Lactate was also positively correlated with hsTNT (Figure 2F). Moreover, PDK4 additionally correlated with ICU length of stay (Figure 2G). Receiver operating characteristic (ROC) curve analysis demonstrated that PDK4 (area under the curve [AUC] = 0.8516) and lactate (AUC = 0.7106) have predictive value for septic myocardial injury (Figure 2H).

**Figure 2.**
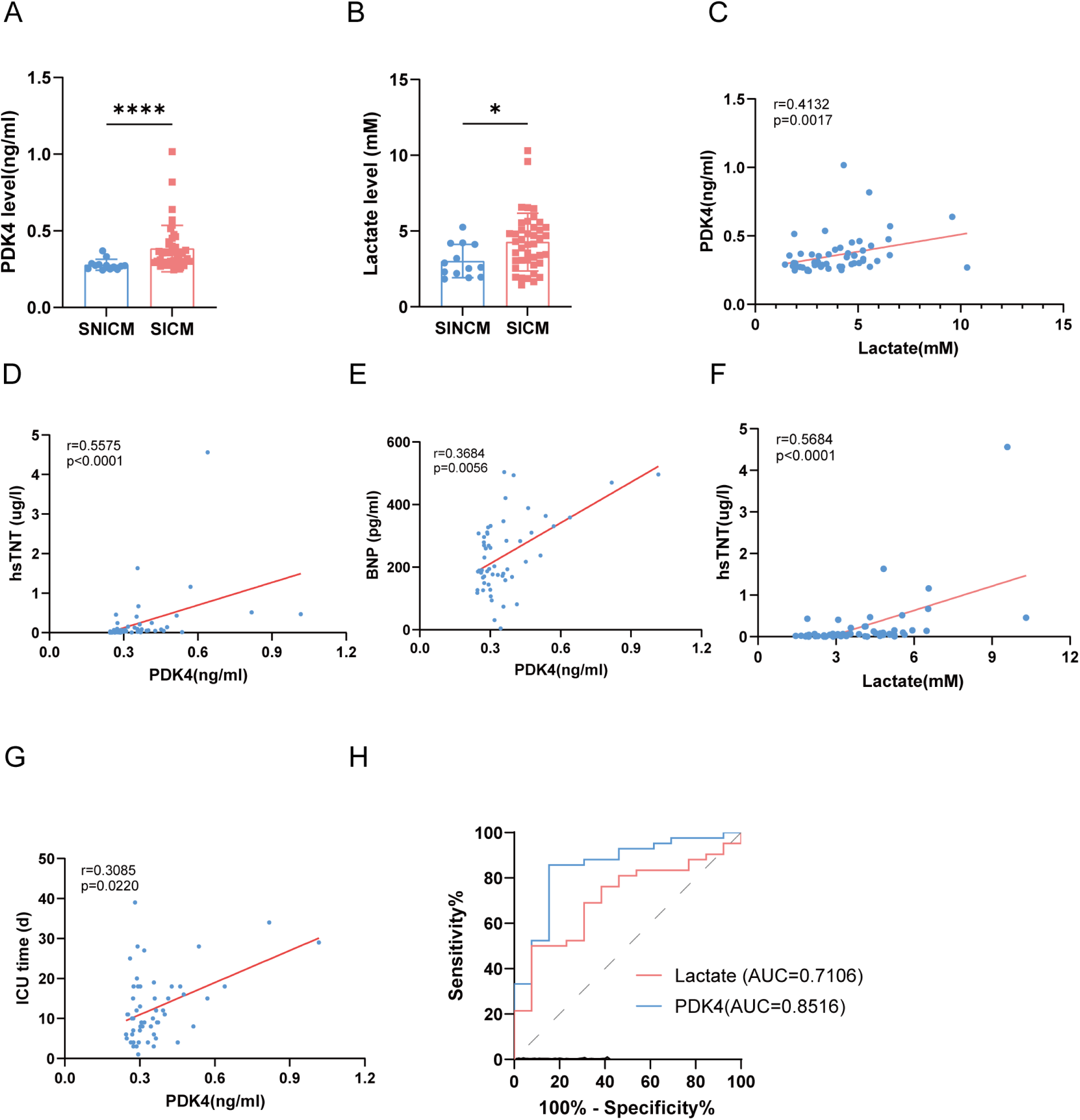
PDK4 and lactate may serve as biomarkers for SICM in patients with sepsis. (A) Serum PDK4 levels in septic patients with or without myocardial injury (SINCM (*n*=13), SICM (*n*=42) (B) Serum lactate levels in septic patients with or without myocardial injury. SINCM (*n*=13), SICM (*n*=42) (C) Correlation analysis between Serum PDK4 and lactate levels in septic patients with or without myocardial injury (*n*=55) (D-E) Correlation analysis of Serum PDK4 and lactate levels with hsTNT and BNP level (F) Correlation analysis of Serum lactate levels with hsTNT (G) Correlation analysis between PDK4 and total ICU length of stay in septic patients (H) ROC curve analysis of the predictive value of PDK4 and lactate for SICM Data are presented as mean ± SD. Statistical significance was determined by Mann-Whitney U test (A, B) or Spearman correlation analysis (C-G). (\**p* < 0.05, \*\**p* < 0.01, \*\*\**p* < 0.001, and \*\*\*\**p* < 0.0001.)

### Lactate accumulation exacerbates SICM

Further analysis revealed a positive correlation between lactate accumulation and the elevation of myocardial injury markers in mice (Figure 3A-3B) consistent with the findings in the human study mentioned above, suggesting lactate may serve as a potential mediator of cardiac damage in SICM, as depicted in the schematic (Figure 3C). To investigate this hypothesis, we employed both gain-of-function and loss-of-function strategies: exogenous lactate (LA) supplementation to mimic pathological hyperlactatemia and sodium oxamate (OX) administration to inhibit lactate dehydrogenase (LDH)-mediated lactate production (Figure 3D).

**Figure 3.**
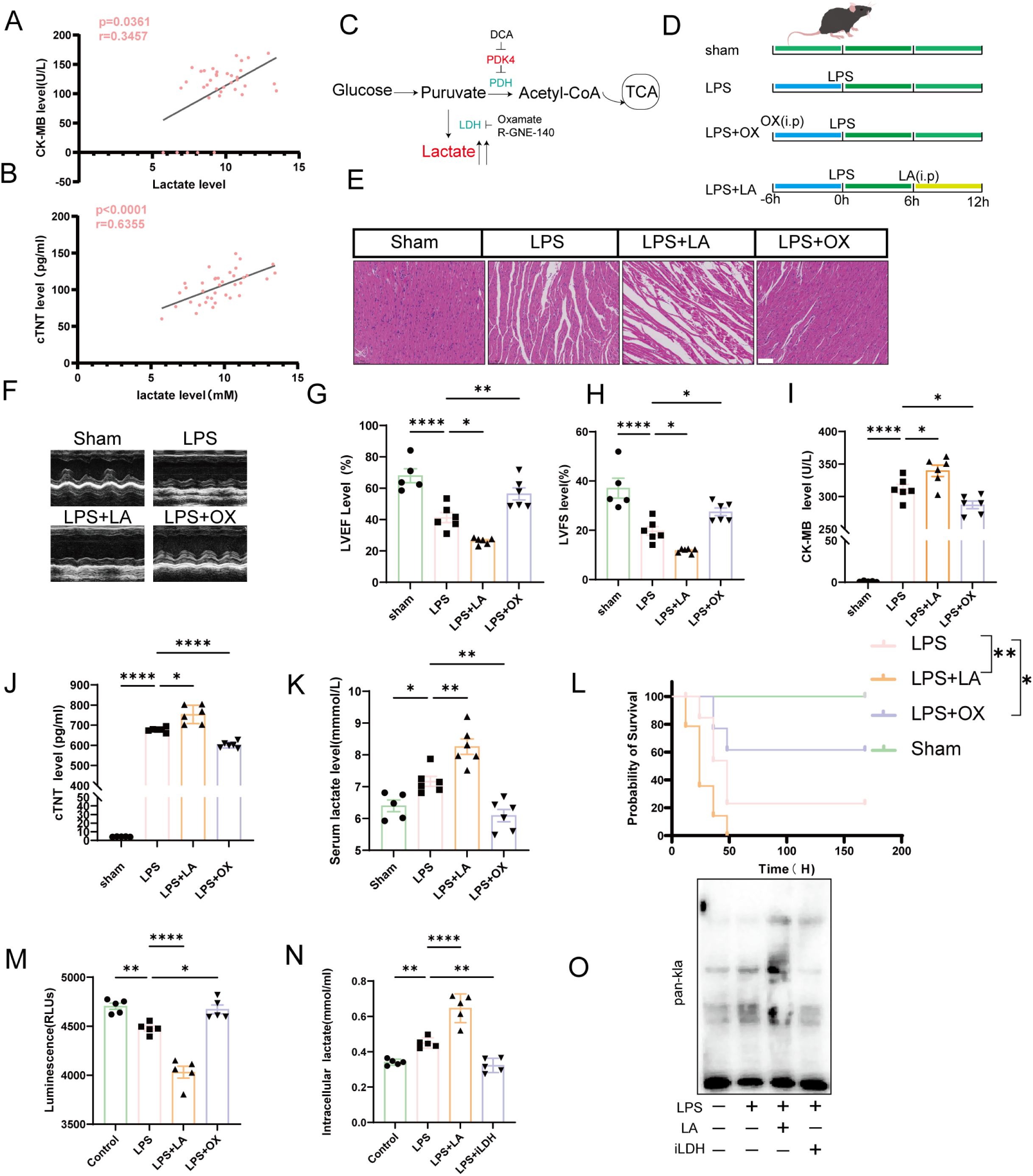
Lactate accumulation exacerbates SICM. (A, B) Correlation analysis between serum lactate concentration and myocardial enzyme levels (CK-MB, cTnT) in mice (n=37). (C) Schematic diagram of cellular energy metabolism pathways, highlighting lactate’s role as a metabolic hub in sepsis. (D) Experimental design schematic: sham group, lipopolysaccharide (LPS)-treated group, LPS combined with sodium oxamate (OX) group, and LPS combined with exogenous lactate group. (E) Representative hematoxylin-eosin (HE) staining images of myocardial tissue (scale bar: 100 μm). (F-H) Representative M-mode echocardiographic images and quantitative analysis of cardiac function parameters (LVEF and LVFS) (sham group, n=5; treated groups, n=6). (I, J) Serum levels of myocardial injury markers (CK-MB, cTnT). (K) Serum lactate concentrations in each group (n=6 per group). (L) Survival curve analysis of different treatment groups (sham, LPS, LPS + lactate, LPS + OX (iLDH); n=14 per group. (M) Viability of H9C2 cardiomyocytes under different treatments (control, LPS, LPS + lactate, LPS + iLDH; n=5 per group). (N) Intracellular lactate content in H9C2 cells (n=5 per group). (O) Western blot analysis of total protein lactylation modification levels in H9C2 cardiomyocytes. The statistical significance of the differences between groups was determined by Spearman correlation analysis (A) and Pearson correlation analysis(B) and one-way ANOVA with Dunnett’s analysis (G,H,I, K, M,N) and Brown-Forsythe and Welch ANOVA tests (J) and Log-rank (Mantel-Cox) test (L) (n.s., not significant; *p < 0.05, **p < 0.01, ***p < 0.001, ****p < 0.0001versus sham or LPS)

Exogenous lactate administration significantly aggravated cardiac dysfunction, as evidenced by decreased LVEF and Left Ventricular Fractional Shortening (LVFS) and increased levels of serum myocardial enzymes compared to LPS-treated controls. In contrast, sodium oxamate treatment attenuated LPS-induced cardiac injury and normalized myocardial injury biomarkers (Figure 3E-3J). Metabolic profiling confirmed these effects, with markedly elevated serum lactate concentrations in the lactate-supplemented group and a substantial reduction following LDH inhibition (Figure 3K). Survival analysis further underscored the detrimental impact of lactate accumulation: the exogenous lactate group exhibited a 100% mortality rate, whereas oxamate treatment significantly improved survival compared to the LPS group (Figure 3L).

Consistent with these *in vivo* findings, *in vitro* experiments using H9C2 cells demonstrated decreased cell viability and elevated intracellular lactate levels upon lactate exposure, while LDH inhibition preserved cell viability and reduced intracellular lactate accumulation compared with the LPS treatment group (Figure 3M-3N). To explore the molecular mechanism whereby lactate causes cardiac damage, we explored protein lactylation, a newly reported lactate-dependent post-translational modification ^16^. Indeed, western blot analysis revealed increased global protein lactylation in the lactate-treated group compared to the LPS treatment group, whereas the LDH inhibition-treated group exhibited reduced levels. (Figure 3O). Collectively, these findings strongly suggest that lactate exacerbates SICM, potentially by promoting protein lactylation.

### PDK4 Induces Lactylation via Lactate Production

Next, we sought to investigate whether lactylation plays a mechanistic role in the pathogenesis of SICM. To address this, we first established an *in vitro* SICM model and analyzed cellular responses over time following LPS stimulation. Using a pan-lactylation antibody, we observed time-dependent increases in protein lactylation levels, with pronounced elevations at 12 and 48 hours post-LPS treatment (Figure 4A). Consistent with these findings, cardiac tissues from LPS-treated SICM mice also exhibited significantly elevated lactylation levels (Figure 4B). Notably, inhibition of PDK4 effectively attenuated this lactylation increase (Figure 4C), further indicating that PDK4 promotes lactylation.

**Figure 4.**
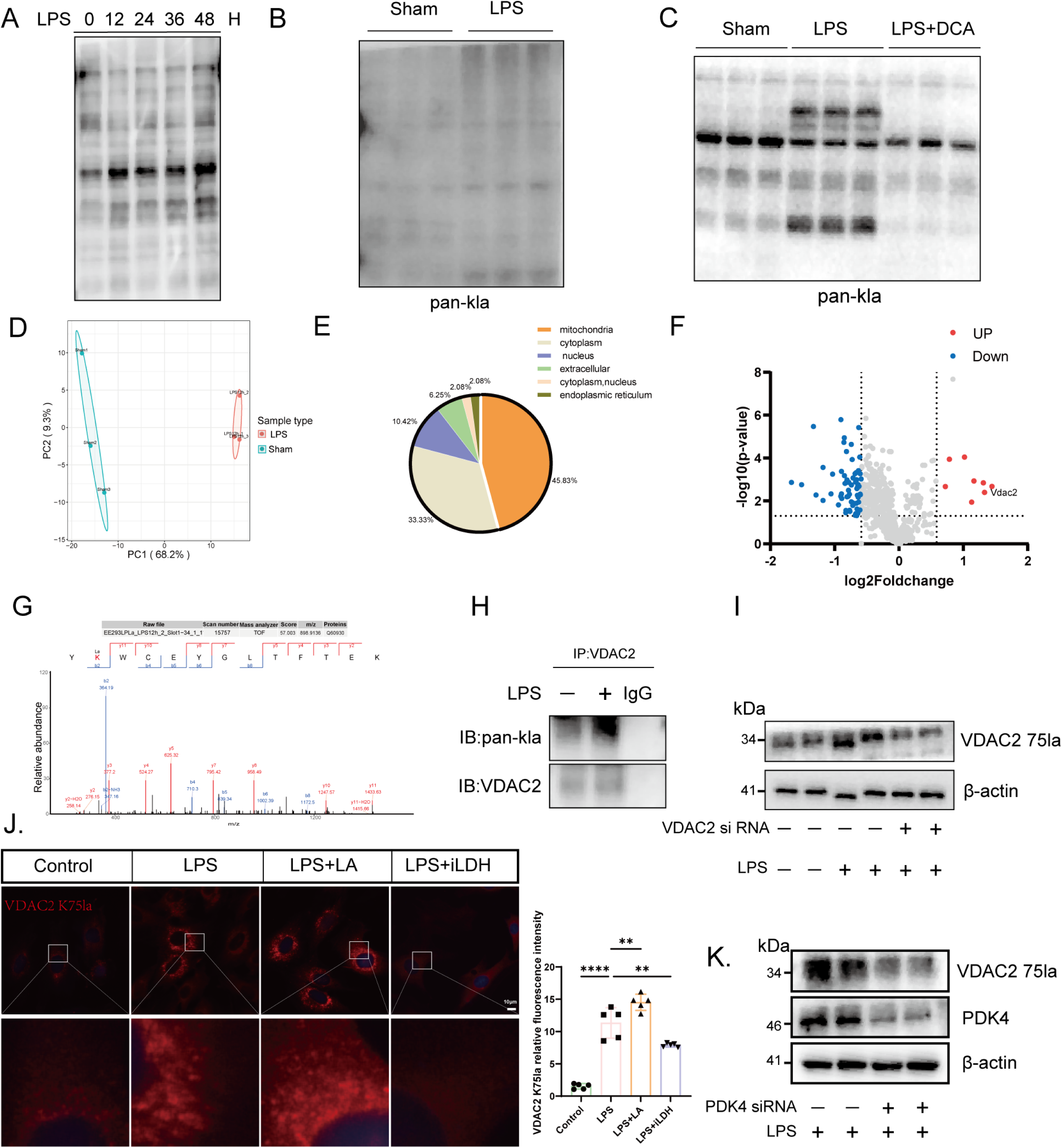
PDK4 Induces Lactylation via Lactate Production. (A) Total protein lactylation levels in H9C2 cardiomyocytes after LPS stimulation at different time points. (B) Total protein lactylation levels in myocardial tissues of mice with or without LPS treatment (*n*=3). (C) Total protein lactylation levels in myocardial tissues of mice after sham, LPS, or LPS combined with dichloroacetate (DCA) treatment (*n*=3). (D) Principal component analysis (PCA) of protein lactylation profiles, showing the first two principal components with centroids linked by sample type (*n*=3). (E) Pie chart depicting the subcellular localization of differentially lactylated proteins in H9C2 cells after 12-hour LPS stimulation. (F) Volcano plot of differentially lactylated proteins in H9C2 cells after 12-hour LPS stimulation. (G) Mass spectrometry abundance profile of lactylated VDAC2 at lysine 75 (K75la) site. (H) Co-immunoprecipitation (Co-IP) showing VDAC2 K75la levels in myocardial tissues with or without LPS treatment. (I) Western blot analysis of VDAC2 K75la levels in H9C2 cells transfected with control (siNC) or *VDAC2*-targeting siRNA (si-*VDAC2*) followed by LPS stimulation. (J) Immunofluorescence staining of VDAC2 K75la in H9C2 cells treated with control, LPS, LPS + lactate, or LPS + lactate dehydrogenase inhibitor (iLDH) (*n*=5). (K) Western blot showing VDAC2 K75la levels in H9C2 cells with or without *PDK4* knockdown followed by 12-hour LPS stimulation. Data are mean ± SD. The statistical significance of the differences between groups was determined by one-way ANOVA with Dunnett’s post hoc analysis (J) (*n.s*, not significant; \**p* < 0.05, \*\**p* < 0.01 versus control or LPS).

To identify key proteins undergoing lactylation, we performed liquid chromatography-tandem mass spectrometry (LC-MS/MS) proteomic analysis on cardiac tissues from LPS-treated mice (Figure 4D, Figure S2). Remarkably, 45.8% of lactylated proteins localized to mitochondria (Fig.4E). This finding prompted us to focus on the mitochondrial outer membrane channel protein voltage-dependent anion channel 2 (VDAC2) (Figure 4F-4G), which regulates cellular Ca²⁺ homeostasis and ferroptosis and is critically involved in cardiac function ^30, 33^. Co-immunoprecipitation assays confirmed significantly increased lactylation of VDAC2 in myocardial tissues from LPS-treated mice (Figure 4H). *In vitro*, LPS stimulation markedly elevated VDAC2 lactylation in H9C2 without altering total VDAC2 protein expression (Figure S3A-S3B), findings further validated using Flag-tagged *VDAC2* plasmid overexpression followed by immunoprecipitation (Figure S3C). Dose-response experiments revealed that 5 µg/ml LPS induced maximal VDAC2 lactylation, which corresponded with the lowest observed cell viability (Figure S3D-S3E). Additionally, an inverse correlation was observed between cell viability and lactate levels, while lactylation levels positively correlated with lactate concentration (Figure S3F-S3G), suggesting that VDAC2 lactylation may contribute to SICM pathogenesis.

To further validate these findings, we generated a custom antibody specific to VDAC2 K75la and used *VDAC2*-targeting siRNA to knock down its expression. This intervention significantly reduced VDAC2 K75la levels (Figure 4I), thereby demonstrating that the customized VDAC2 K75la antibody specifically recognizes the VDAC2 lactylated peptide. Immunofluorescence staining and a western blot revealed heightened VDAC2 K75la signals in lactate-treated cardiomyocytes compared to LPS-only treated cells, whereas LDH inhibition significantly diminished these signals (Figure 4J, S3H). Cell viability assays further demonstrated that *VDAC2* knockdown protected against LPS-induced injury, an effect reversed by exogenous lactate treatment, indicating that lactate exerts its effects via VDAC2 K75la (Figure S3I).

To investigate PDK4’s upstream role, we engineered *PDK4*-specific siRNA and overexpression plasmids. *PDK4* knockdown significantly reduced VDAC2 lactylation levels (Figure 4K), while overexpression increased them (Figure S4A). Critically, inhibiting glycolysis with 2-DG following *PDK4* overexpression reversed VDAC2 lactylation (Figure S4A), indicating PDK4 promotes lactylation via glycolysis. Functionally, exogenous lactate compromised cell viability in NC controls, whereas *PDK4* knockdown restored viability (Figure S4B). Consistent with these findings, lactate exacerbated cardiac dysfunction and elevated serum cardiac enzymes in septic mice—effects that were reversed by DCA co-administration (Figure S4C-S4D).

In summary, these findings establish that PDK4 drives VDAC2 K75la via glycolysis. This novel mechanism reveals a critical role for post-translational modification in the metabolic regulation of SICM and highlights PDK4 as a promising therapeutic target.

### VDAC2 lactylation exacerbates SICM, with lysine 75 (K75) identified as the critical modification site

Our findings strongly support the pivotal role of VDAC2 K75la in the pathogenesis of SICM. Lactomics analysis identified a significant increase in VDAC2 K75la. Evolutionary conservation analysis further revealed that this lysine residue is relatively well-conserved across multiple species such as humans, mice, and gorillas (Figure 5A), underscoring its potential functional importance. To investigate the specific contribution of VDAC2 K75la, we engineered two adeno-associated viral (AAV) constructs: AAV-FLAG-VDAC2 (K75) (wild-type, WT) and AAV-FLAG-VDAC2 (R75) (arginine-substitution mutant, Mut), the latter being a mutant that prevents lactylation at K75 of VDAC2 (Figure 5B). These constructs were administered via tail vein four weeks before LPS-induced sepsis modeling (Figure 5C).

**Figure 5.**
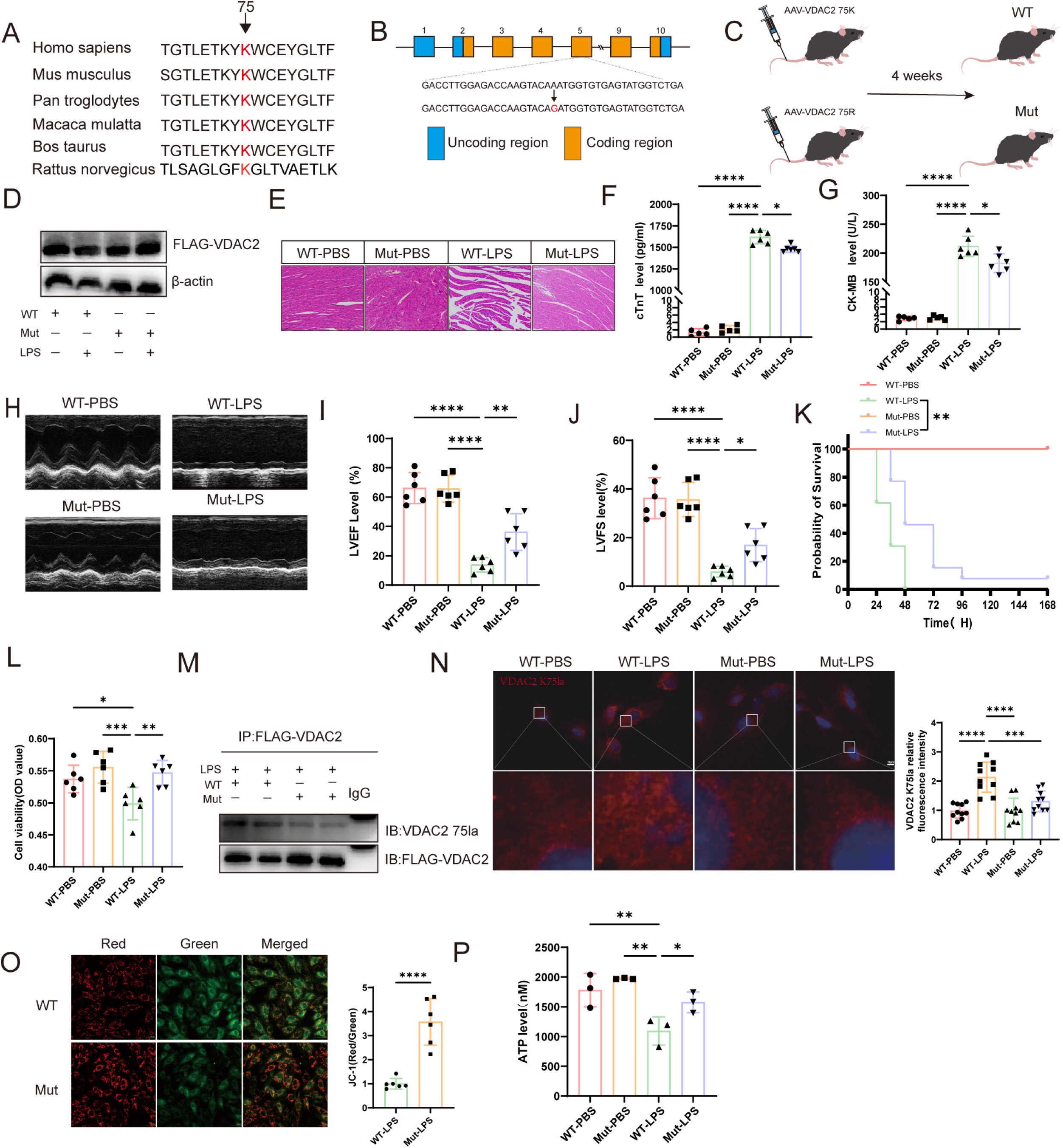
VDAC2 lactylation exacerbates SICM, with lysine 75 (K75) identified as the critical modification site. (A) Multiple sequence alignment analysis of the VDAC2 K75 residue across species. (B) Native VDAC2 structure (top) and mutant VDAC2 structure (bottom) highlighting the K75 mutation site. (C) Experimental design schematic: Mice injected with AAV-VDAC2-WT or AAV-VDAC2-Mut treated with PBS or LPS via intraperitoneal injection. (D) Western blot analysis of FLAG-VDAC2 expression in myocardial tissues of mice injected with AAV-WT or AAV-Mut constructs. (E) Representative H&E-stained images of myocardial tissues (scale bars: 100 μm). (F-G) Serum myocardial injury marker (CK-MB, cTnT) levels in experimental groups (*n*=6). (H-J) Representative M-mode echocardiograms and Echocardiographic parameters (LVEF, LVFS). (K) Survival analysis of WT-PBS (*n*=6), WT-LPS (*n*=13), Mut-PBS (*n*=6), and Mut-LPS (*n*=13) groups. (L) Viability of H9C2 cells from WT and Mut groups with or without LPS treatment (*n*=8). (M) Co-IP with anti-Flag antibody followed by Western blot with anti-Kla antibody in H9C2 cells transfected with Flag-*VDAC2*-WT or Falg-*VDAC2*-Mut plasmids in H9C2 cells (*n*=3). (N) Representative immunofluorescence images of VDAC2 K75la in H9C2 cardiomyocytes (scale bars: 10 μm). (O) Mitochondrial membrane potential in WT and Mut cardiomyocytes (*n*=5). (P) ATP content in WT and Mut H9C2 cardiomyocytes with or without LPS treatment (*n*=3). The statistical significance of the differences between groups was determined by and Brown-Forsythe and Welch ANOVA tests (F, J) and one-way ANOVA with Dunnett’s analysis analysis (H,I,P) and Log-rank (Mantel-Cox) test (K) and unpaired Student’s t test (O) (*n.s.*, not significant; \**p* < 0.05, \*\**p* < 0.01, \*\*\**p* < 0.001, \*\*\*\**p* < 0.0001versus WT-LPS)

Western blot confirmed consistent FLAG-VDAC2 expression levels in both WT and Mut groups (Figure 5D). Histopathological examination showed that mice expressing the Mut construct exhibited reduced myocardial fiber disruption and less interstitial edema compared to the WT group (Figure 5E). Correspondingly, biochemical analyses revealed lower levels of cardiac injury biomarkers (Figure 5F-5G) and significantly improved cardiac function in the Mut group, as evidenced by enhanced LVEF and LVFS (Figure 5H-5J). Notably, Mut mice demonstrated a significantly higher survival rate following sepsis induction (Figure 5K).

Consistent with *in vivo* findings, *in vitro* experiments using H9C2 cardiomyocytes transfected with WT or Mut plasmids demonstrated similar trends. Following transfection, the plasmid showed marked RNA-level upregulation, confirming stable intracellular expression (Figure S5A). After LPS stimulation, WT-expressing cells showed significantly reduced viability compared to the Mut group (Figure 5L, Figure S5B). Immunoprecipitation (IP) and immunofluorescence imaging revealed higher VDAC2 K75la signal intensity in WT cells (Figure 5M-5N).

Mitochondrial function assays revealed marked dysfunction in WT cells, including significantly reduced mitochondrial membrane potential (Figure 5O) and lower ATP production (Figure 5P). Moreover, WT cells accumulated higher levels of reactive oxygen species (ROS) following LPS treatment, compared to the Mut group (Figure S5C). Given that VDAC2 modulates intracellular Ca²⁺ handling, we measured cytosolic Ca²⁺ levels and found that LPS stimulation elicited a significantly greater increase in WT cells than in Mut cells, suggesting that VDAC2 lactylation may impair its intrinsic Ca²⁺-regulatory function (Figure S5D), consistent with previous studies suggesting that PDK4 can influence intracellular calcium dynamics ^34^. This finding suggest that PDK4 may mediate calcium dysregulation in SICM via VDAC2 K75la. Furthermore, we examined the effects of exogenous lactate on mitochondrial function, which produced results consistent with those observed in WT-expressing cells, reinforcing the pathogenic role of lactate-driven VDAC2 K75la (Figure S5E-S5F).

Taken together, these comprehensive findings demonstrate that VDAC2 K75la constitutes a critical regulatory modification through which VDAC2 mediates metabolic dysfunction and myocardial injury in SICM. This study offers novel mechanistic insights into the molecular underpinnings of SICM and identifies VDAC2 K75 lactylation as a potential therapeutic target.

### VDAC2 K75la Exacerbates SICM by Inhibiting Autophagic Flux through Interaction with NBR1

The above findings demonstrate that VDAC2 K75la exacerbates SICM. To elucidate the underlying mechanism, we performed co-immunoprecipitation (co-IP) with mass spectrometry, which detected a reduced interaction between the WT group and NBR1 compared to the Mut group (Figure 6A and Figure S6A). Silver staining performed after co-IP of proteins also showed that the protein in the WT group was far less common at the 140 kDa position, as corroborated with the observation (Figure S6B). Subsequent co-IP validation confirmed weakened NBR1-VDAC2 binding in the WT group relative to the Mut group (Figure 6B). Our computational modeling has identified putative interaction domains between VDAC2 and NBR1, with molecular docking simulations revealing a high-probability binding interface (Figure S6C). Immunofluorescence co-localization assays further supported these results, showing enhanced NBR1-VDAC2 co-localization in Mut cells (Figure 6C).

**Figure 6.**
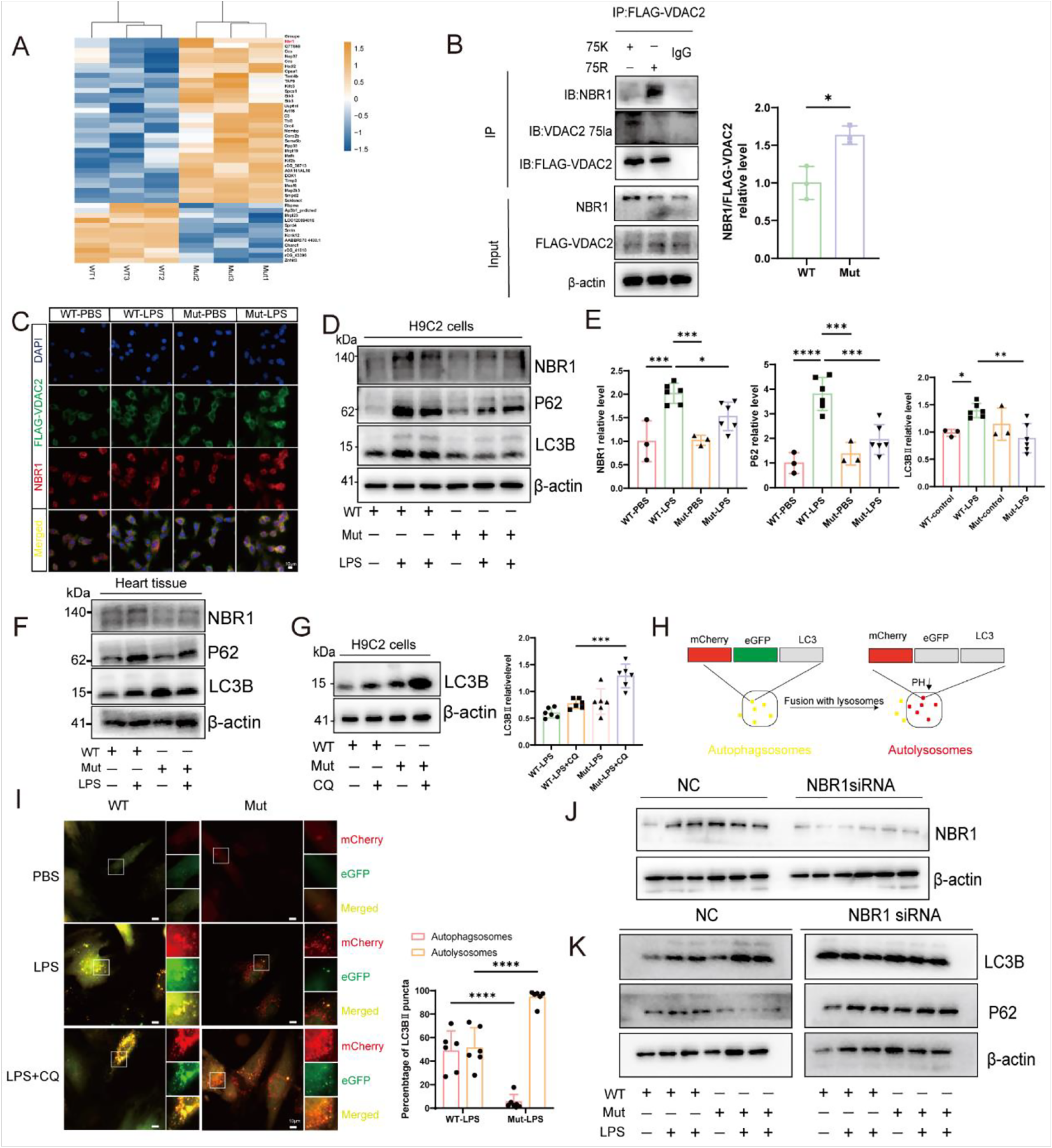
VDAC2 K75la Exacerbates SICM by Inhibiting Autophagic Flux through Interaction with NBR1. (A) Heatmap of proteins identified by co-immunoprecipitation (CO-IP) mass spectrometry in wild-type (WT) and mutant (Mut) H9C2 cardiomyocytes (*n*=3). (B) Co-IP assays demonstrated that VDAC2 directly interacts with NBR1 in WT and Mut H9C2 cells (*n*=3). (C) Dual immunofluorescence staining of VDAC2 K75la and NBR1, showing their co-localization in WT-LPS (*n*=7), WT-PBS (*n*=7), and Mut-LPS (*n*=9) groups (scale bars: 10 μm). (D, E) Western blot analysis and quantification of autophagy-related proteins (P62, NBR1, LC3B) in H9C2 cells and myocardial tissues from WT and Mut groups with or without LPS treatment. (F) Expression levels of P62, NBR1, and LC3B in myocardial tissues of WT and Mut mice. (G) Western blot showing LC3B levels in WT and Mut groups after LPS treatment with or without chloroquine (CQ). (H) Schematic diagram of the LC3 dual-fluorescence reporter system. (I) Representative co-localization images of the LC3 dual-fluorescence assay (*n*=6; scale bars: 10 μm). (J) Western blot showing NBR1 levels in H9C2 cells with or without *NBR1* siRNA knockdown (*n*=6). (K) Western blot analysis of P62 and LC3B expression in H9C2 cells with or without NBR1 siRNA-mediated *NBR1* knockdown. The statistical significance of the differences between groups was determined by unpaired two-tailed Student’s t test(B) or one-way ANOVA with Dunnett’s analysis (E) and Brown-Forsythe and Welch ANOVA tests (C) and Two-way ANOVA with Tukey’s multiple comparisons test(I) (*n.s.*, not significant; \**p* < 0.05, \*\**p* < 0.01, \*\*\**p* < 0.001, \*\*\*\**p* < 0.0001versus control, WT-LPS or Mut-LPS)

Given the role of NBR1 in selective autophagy, we hypothesized that VDAC2 K75la modulates autophagic processes. Western blot analysis of key autophagy markers revealed significant accumulation of p62 and NBR1 in WT cells compared to the Mut group, alongside elevated LC3-II levels in the WT group (Figure 6D-6E). A parallel trend was observed in murine myocardial tissue (Figure 6F). These data collectively suggest that VDAC2 K75la impairs autophagic flux, suppressing autophagy and exacerbating myocardial injury.

To directly assess autophagic flux, we inhibited lysosomal degradation using chloroquine (CQ). Following CQ treatment, WT cells exhibited fewer LC3-II levelsthan the Mut group (Figure 6G), confirming impaired autophagosome-lysosome fusion in the WT group. LC3 dual-fluorescence assays further illustrated autophagic dynamics: WT cells showed predominant LC3-II accumulation, whereas Mut cells displayed increased autolysosome formation (Figure 6H-6I), indicating significantly impairment of autophagic flux induced by VDAC2 K75la. Next, we constructed *NBR1* siRNA. Upon *NBR1* knockdown, Western blot analysis revealed no significant differences in autophagy-related protein levels between WT and Mut groups. However, in the NC group, autophagic flux was significantly inhibited in the WT group (Figure 6J-6K), suggesting that VDAC2 may mediate its effects through NBR1.

We also conducted *VDAC2* knockdown and overexpression experiments. *VDAC2* knockdown reduced the accumulation of p62 and NBR1 and enhanced autophagy compared to the NC group (Figure S7B-S7C). After CQ treatment, the LC3B-II level was higher in the *VDAC2* knockdown group than in the control (Figure S7D), suggesting impaired autophagosome-lysosome fusion in the NC group, leading to blocked autophagy. Conversely, *VDAC2* overexpression yielded opposite effects (Figure S7E-S7G). Additionally, myocardial enzyme levels in the supernatant of H9C2 cells were lower following *VDAC2* knockdown compared to the NC group (Figure S7H), further supporting the notion that VDAC2 lactylation impairs autophagic flux and aggravates SICM. These findings also suggest that *VDAC2* knockdown mitigates myocardial injury. To further validate that lactate acts via VDAC2 lactylation, we supplemented exogenous lactate. In the NC group, lactate significantly increased p62 accumulation. In contrast, p62 levels were markedly reduced after *VDAC2* knockdown, indicating that lactate impairs autophagy by promoting VDAC2 lactylation, which exacerbates cell death (Figure S7I). To further confirm that VDAC2 K75la suppresses autophagic flux, we employed *Pdk4* knockout in mice and *PDK4* knockdown in H9C2 cells. *PDK4* knockdown significantly reduced VDAC2 K75la levels, decreased p62 and NBR1 accumulation, and increased LC3-II levels. These results indicate that *PDK4* knockdown not only promotes autophagy initiation but also enhances autophagic flux (Figure S8).

In conclusion, our comprehensive findings demonstrate that VDAC2 K75la lactylation exacerbates SICM by inhibiting autophagic flux, providing novel insights into the molecular mechanisms underlying SICM.

### CBP is the writer promoting VDAC2 lactylation

Considering the role of VDAC2 K75la in SICM, we attempted to identify its writer. We first examined the conventional writers CREB-binding protein (CBP)/P300 and used siRNA to knock down their expression (Figure 7A). We found that knockdown of CBP/P300 significantly reduced the level of VDAC2 K75la (Figure 7B). Notably, our quantitative proteomic analysis revealed a marked upregulation of CBP following LPS stimulation (Figure 7C). Additionally, after CBP knockdown, the levels of cTNT in the cell supernatant were lower compared to the NC group (Figure 7D). Furthermore, through IP, we discovered an interaction between VDAC2 and CBP, with higher lactylation levels of VDAC2 correlating with stronger interactions with CBP (Figure 7E). These results suggest that CBP is likely the writer for VDAC2 lactylation.

**Figure 7.**
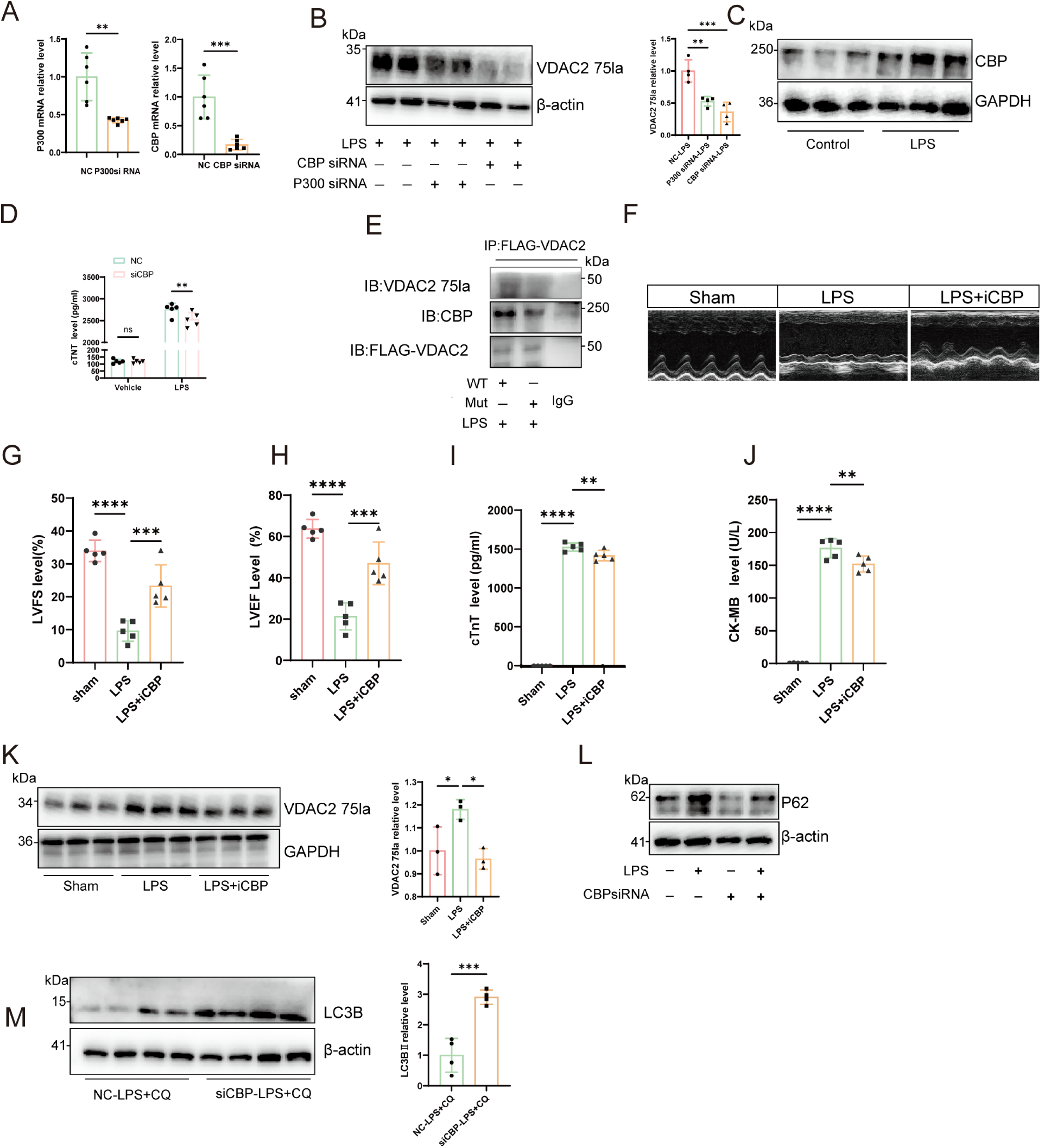
CBP is the writer promoting VDAC2 lactylation. (A) mRNA levels of P300 and CBP in H9C2 cells after siRNA-mediated knockdown of P300 or CBP (*n*=6). (B) Western blot analysis of VDAC2 K75la levels in H9C2 cells following P300 or CBP knockdown (*n*=4). (C) Western blot showing CBP expression in H9C2 cells with or without LPS stimulation (*n*=3). (D) cTnT levels in cell supernatants with or without CBP knockdown and LPS treatment (*n*=5). (E) Western blot demonstrating interaction between VDAC2 and CBP. (F-H) Representative echocardiograms and cardiac function parameters in Sham, LPS, and LPS+iCBP-treated groups (*n*=5). (I-J) Serum cardiac enzyme (CK-MB, cTnT) levels in Sham, LPS, and LPS+iCBP-treated mice (*n*=5). (K) Western blot analysis of VDAC2 K75la levels in myocardial tissues from Sham, LPS, and LPS+iCBP groups (*n*=3). (L) Western blot showing P62 expression in H9C2 cells with or without CBP knockdown and LPS treatment (*n*=3). (M) Western blot analysis of LC3B levels in H9C2 cells with CBP knockdown, LPS stimulation, and chloroquine (CQ) treatment (*n*=4). The statistical significance of the differences between groups was determined by unpaired two-tailed Student’s *t*-test (A, M) and one-way ANOVA with Dunnett’s analysis(H-K) and Two-way ANOVA with Bonferroni’s multiple comparisons test (D) (*n.s.*, not significant; \**p* < 0.05, \*\**p* < 0.01, \*\*\**p* < 0.001, \*\*\*\**p* < 0.0001versus LPS)

**Figure 8.** Mechanism Diagram of PDK4-VDAC2 Lactylation-Impaired Autophagy Axis in Regulating Sepsis-Induced Myocardial Injury. PDK4, located in the mitochondrial matrix, is upregulated during SICM. It enhances glycolysis in cardiomyocytes, leading to further accumulation of lactate. The increased lactate promotes lactylation of VDAC2, specifically reducing its interaction with NBR1, thereby impairing autophagy and exacerbating myocardial damage.

To explore the role of CBP in regulating VDAC2 lactylation during SICM, we administered a CBP inhibitor to mice via gavage for one week before inducing sepsis with LPS. We found that in the sepsis myocardial injury model treated with the CBP inhibitor (iCBP), cardiac function was significantly improved, and levels of myocardial injury biomarkers were reduced (Figure 7F-7J). Western blot analysis also revealed decreased lactylation levels of VDAC2 in myocardial tissue from iCBP mice (Figure 7K). Moreover, after CBP knockdown, we observed significant reductions in p62 levels in H9C2 cells (Figure 7L), and using chloroquine, accumulation of LC3B-II was pronounced in CBP-knockdown cardiomyocytes, compared to the modest elevation in control cells (Figure 7M), indicating that inhibiting CBP promotes autophagy, thereby alleviating SICM.

Overall, these results demonstrate that CBP is likely the writer for VDAC2 K75 lactylation, and inhibition of CBP may represent a promising therapeutic target for effectively alleviating SICM.

## Discussion

In this study, *PDK4* upregulation drives enhanced glycolysis and subsequent lactate accumulation. Notably, this accumulated lactate acts not merely as a metabolic byproduct, but as a signaling molecule that specifically induces VDAC2 lactylation at the K75 site. This pathological modification disrupts VDAC2-NBR1 binding, impairing selective autophagy flux in cardiomyocytes. Consequently, toxic substrates accumulate, exacerbating SICM. Our study establishes the first direct molecular connection between metabolic reprogramming and autophagy dysfunction in sepsis, highlighting the critical role of protein lactylation in SICM pathophysiology. These findings not only provide novel insights into SICM pathogenesis but also identify promising therapeutic targets.

During the process of SICM, lactate levels are elevated. Traditionally, this hyperlactatemia was attributed to tissue hypoxia. However, in SICM, myocardial tissue rarely experiences true oxygen deficiency; in fact, with appropriate resuscitation, cardiac oxygenation can even exceed normal levels ^35^. This suggests that elevated lactate in SICM stems primarily from metabolic reprogramming rather than hypoxia. Significant metabolic reprogramming during SICM is well-documented. For instance, PKM2 inhibition effectively suppresses glycolysis and reduces inflammatory cytokine release ^10^. Similarly, KLF14 decreases macrophage glycolysis and inflammatory cytokine secretion by inhibiting *HK2* transcription ^11^. Moreover, inhibiting PDK4 has been suggested to alleviate SICM, although the precise mechanisms remain unclear ^15^. Our research demonstrates that PDK4 exacerbates SICM by promoting lactate accumulation through glycolytic regulation. Specifically, PDK4 inhibits pyruvate dehydrogenase, diverting pyruvate away from the TCA cycle and toward lactate production ^13, 36^. Although we also observed upregulation of PFKFB3, a key regulator of glycolysis, we targeted PDK4 since it directly controls the fate of pyruvate, the immediate precursor of lactate, and thus represents a key node linking glycolysis to the lactate-dependent signaling that we wished to investigate. This study provides compelling evidence that PDK4-driven upregulation of glycolysis is a key mechanism underlying non-hypoxic lactate accumulation in SICM. Moreover, our results show that these elevated lactate levels directly contribute to myocardial injury, indicating lactate acts pathogenically not only as a metabolic marker but also as a mediator of cardiac damage. Supporting these findings, detection of serum PDK4 levels in septic patients revealed positive correlations between PDK4 and lactate levels, cardiac injury markers, and ICU length of stay. Therefore, targeting PDK4 or lactate production represents a promising therapeutic strategy to mitigate septic cardiomyopathy.

Once considered merely a metabolic byproduct, lactate has undergone a paradigm shift in its biological significance. Emerging research now recognizes lactate as a critical signaling molecule with diverse roles in immune regulation ^17^, signal transduction ^37^, and cell death ^38^. This functional versatility is partly attributed to its role as a substrate for post-translational modification via protein lactylation ^16^.Lactate has emerged as a valuable prognostic indicator in patients with sepsis ^39, 40^. Recent studies reveal that lactate-driven HMGB1 lactylation and acetylation compromise endothelial integrity by downregulating cadherin and claudin-5 ^41^. Additionally, H3K18la-mediated activation of *Egr1* exacerbates pulmonary vascular leakage ^20^. The pathogenic role of lactylation extends to renal involvement in sepsis, where RHOA-driven inflammation contributes to acute kidney injury ^22^. Collectively, these findings establish lactylation as a key regulator of multi-organ dysfunction in sepsis. Notably, current studies indicate that HADHA lactylation can promote myocardial suppression ^42^, but the role of protein lactylation in SICM has not yet been comprehensively investigated. Our study builds on this concept by identifying VDAC2 K75 lactylation as a pivotal pathogenic event in SICM. Furthermore, research shows that lactylation exacerbates cardiac fibrosis via TGF-β signaling activation in advanced acute myocardial infarction (AMI) stages, yet paradoxically promotes cardiac repair through monocyte activation in early stages ^43, 44^. These contrasting effects highlight the complex and context-dependent roles of lactylation in cardiovascular disease. Its therapeutic potential is intricately linked to factors such as disease stage, cell type, and the specific targets of lactylation, emphasizing the need for precision in therapeutic strategies targeting this modification. We demonstrate that a PDK4-mediated glycolytic shift increases lactate levels, and that both endogenous and exogenous lactate sources potentiate VDAC2 K75la modification. This discovery provides a molecular explanation for the clinical link between hyperlactatemia and poor outcomes in septic patients. In addition, previous studies have demonstrated that PDK4 can induce Ca²⁺ overload in H9C2 cells ^34^. Our findings further suggest that lactylation of VDAC2 may impair its normal function, resulting in dysregulated Ca²⁺ transport and subsequent cellular Ca²⁺ overload, which may contribute to cellular dysfunction. This mechanism may also help explain how elevated PDK4 levels trigger Ca²⁺ overload. Therefore, inhibiting VDAC2 lactylation is a promising therapeutic avenue, and thus the development of targeted drugs against VDAC2 lactylation warrants further investigation as a potential treatment strategy.

One of the key findings of our study is how VDAC2 lactylation leads to dysfunction in the cell. We defined the link between downstream VDAC2 K75 lactylation effects—mitochondrial dysfunction/Ca²⁺ overload and impaired autophagic flux. Our findings define a direct molecular mechanism for the suppression of autophagy: VDAC2 K75la reduces its binding interaction with the autophagy receptor NBR1. However, such disease phenotypes are likely to be interdependent but not independent. The chronic cytoplasmic Ca²⁺ overload that we found may well inhibit autophagic flux, potentially by activating calpains that degrade critical autophagy proteins. Therefore, lactylation of VDAC2 can lead to a ‘dual-hit’ injury of cardiomyocytes: it directly inhibits mitophagy cargo recognition through disruption of the VDAC2-NBR1 interaction, and simultaneously creates an autophagy-inhibitory cellular environment through Ca²⁺ dyshomeostasis. Deciphering the major drivers versus amplifying factors in this network is a pressing direction for future studies. We showed that VDAC2 K75la suppresses autophagy by reducing its interaction with NBR1, a novel regulatory mechanism of particular importance in sepsis. Our study revealed that lactylation of VDAC2 significantly reduces its interaction with NBR1, and this weakened binding leads to impaired autophagic flux, manifested by the accumulation of substrates (e.g., p62 and NBR1) (Figure 6D). NBR1 knockdown experiments further confirmed that VDAC2 likely regulates autophagy through NBR1(Figure 6J-6K). Moreover, this study identified CBP as a writer enzyme for VDAC2 K75la, demonstrating that CBP promotes VDAC2 lactylation. Inhibition or knockdown of *CBP* effectively restored autophagic flux, thereby alleviating SICM (Figure 7). These findings validate the authenticity and significance of the VDAC2 K75la pathway and provide an enzymatic intervention target. However, while CBP inhibitors are an effective proof-of-concept, their nonspecificity for global acetylation and lactylation is a cause for concern regarding off-target effects. More selective inhibitors of VDAC2 lactylation may have greater therapeutic promise. The precise molecular mechanism underlying lactylation-mediated reduction in VDAC2-NBR1 interaction remains to be fully elucidated. Future investigations should address whether this modification induces structural conformational changes in VDAC2, alters subcellular localization, or affects co-factor recruitment essential for NBR1 binding. Therefore, by targeting the reduction of VDAC2 lactylation and the therapeutic activation of autophagy, our study offers a promising strategy for the prevention and treatment of SICM.

This study has several limitations. Foremost is the lack of validation in human myocardial tissue affected by SICM, primarily due to ethical and clinical constraints. Cardiac biopsies in septic patients are highly invasive and rarely warranted. Additionally, the acute and unpredictable nature of sepsis limits the feasibility of timely tissue collection, while postmortem samples may not accurately capture dynamic disease processes. Furthermore, our clinical results showed that age, one of the principal cardiovascular risk factors, differed substantially between the SICM and non-SICM cohorts, potentially acting as a confounding variable. Moreover, the LPS-induced mouse model, although widely used, does not fully recapitulate the complexity of human sepsis. Differences in immune responses, metabolic regulation, and disease progression between species may limit the translational relevance of our findings. Similarly, our *in vitro* experiments used the H9C2 cell line, which is valuable but unlikely to fully mimic primary cardiomyocyte responses. Mechanistic questions also remain unresolved—for instance, how VDAC2 lactylation alters its conformation or charge to impair NBR1 binding, and whether this modification influences other VDAC2 functions, such as channel activity, calcium transport, or interactions with mitochondrial proteins. Our finding that VDAC2 lactylation exacerbates Ca²⁺ overload provides an intriguing parallel mechanism, but how it connects to autophagy failure— as a cause or consequence—is to be investigated. Clarifying these aspects is critical to fully understand the therapeutic potential of targeting VDAC2 lactylation. Finally, the therapeutic viability of this target must be rigorously evaluated in terms of efficacy, safety, and potential off-target effects across various preclinical models and long-term studies. Given that CBP inhibitors may broadly affect protein acetylation and lactylation, their specificity and selectivity require thorough investigation to minimize systemic side effects.

In conclusion, our study identifies VDAC2 K75la as a critical metabolic mediator linking elevated glycolytic flux to impaired autophagy in SICM. These findings provide important mechanistic insights into the metabolic dysregulation underlying sepsis. Furthermore, our results suggest that strategies aimed at reducing lactate accumulation or specifically inhibiting VDAC2 lactylation may help restore autophagic flux, offering a potential therapeutic approach for the treatment of SICM.

## Data Availability

All data generated or analyzed during this study are included in this article

## Acknowledgements

None.

## Availability of data and materials

All data generated or analyzed during this study are included in this article.

## Abbreviations

PDK4: Pyruvate Dehydrogenase Kinase 4
VDAC2: Voltage-Dependent Anion Channel 2
NBR1: Neighbor of BRCA1 Gene 1
SICM: Sepsis-Induced Cardiomyopathy
LPS: Lipopolysaccharide
HIF-1α: Hypoxia-Inducible Factor 1-alpha
PKM2: Pyruvate Kinase M2 Isoform
HK2: Hexokinase 2
PDC: Pyruvate Dehydrogenase Complex
LDHA: Lactate Dehydrogenase A
PFKFB3: 6-Phosphofructo-2-Kinase/Fructose-2,6-Biphosphatase 3
DCA: Dichloroacetate
AAV: Adeno-Associated Virus
LVEF: Left Ventricular Ejection Fraction
LVFS: Left Ventricular Fractional Shortening
cTNT: Cardiac Troponin T
CK-MB: Creatine Kinase-MB Isoform
ECAR: Extracellular Acidification Rate
SINCM: Sepsis-Induced Noncompaction Cardiomyopathy
hs-TNT: High-Sensitivity Troponin T
BNP: B-Type Natriuretic Peptide
ROC: Receiver Operating Characteristic
AUC: Area Under the Curve
VDAC2: K75la VDAC2 Lysine 75 Lactylation
Mut: Mutant
CQ: Chloroquine
CBP: CREB-Binding Protein
CO-IP: Co-Immunoprecipitation
iCBP: Inhibitor of CBP

## Supplemental Material

Figure S1-S8

Tables S1–S3

Supplemental Methods

Major Resources Table

